# Executive functioning in Body Dysmorphic Disorder and Obsessive-Compulsive Disorder

**DOI:** 10.1101/2021.04.29.21256321

**Authors:** Long Long Chen, Oskar Flygare, John Wallert, Jesper Enander, Volen Z. Ivanov, Christian Rück, Diana Djurfeldt

## Abstract

**Objective:** To assess executive functions in patients with Body Dysmorphic Disorder (BDD) and Obsessive-Compulsive Disorder (OCD) compared with healthy controls.

**Methods:** Adults diagnosed with BDD (n=26) or OCD (n=29) according to DSM-5, and healthy controls (n=28) underwent validated and computerized neuropsychological tests; spatial working memory (SWM), Intra- extra dimensional set shifting (IED) and Stop signal task (SST), from the Cambridge Neuropsychological Test Automated Battery (CANTAB). Test performance was compared between groups, and correlated to standardized symptom severity of BDD and OCD. Significance level was set to p<0.05.

**Results:** There were no statistically significant between-group differences on key outcome measures in SWM, IED, or SST. There was a weak positive correlation between symptom severity and test errors on SWM and IED in both OCD and BDD groups; increased clinical severity were associated with more errors in these tests. Further, there was a negative correlation between symptom severity and SST in the BDD group.

**Conclusions:** Patients with BDD or OCD did not differ from healthy control subjects in terms of test performance, however there were several statistically significant correlations between symptom severity and performance in those with BDD or OCD. More studies on EF in BDD and OCD are required to elucidate if there are differences in EF between these two disorders.

## INTRODUCTION

Obsessive-Compulsive Disorder (OCD) and Body Dysmorphic Disorder (BDD) share characteristics such as intrusive thoughts and repetitive behaviors (1). Patients with OCD typically experience obsessional thoughts, images or impulses that cause anxiety or distress, which can be neutralized by overt compulsive behaviors or mental acts (2). Patients with BDD are preoccupied with a perceived defect in physical appearance and engage in repetitive excessive grooming, mirror checking or reassurance-seeking behaviors (3).

OCD and BDD share a number of important clinical features, including sex ratio, age of onset and treatment response to selective serotonin reuptake inhibitor (4). Moreover, evidence from a twin study supports a genetic overlap between OCD and BDD (5). Moreover, studies suggest that 27.5% of patients primarily diagnosed with BDD have comorbid OCD, whereas 10.4% of patients primarily diagnosed with OCD have comorbid BDD (6, 7). Although OCD and BDD are presumably related to each other regarding underlying psychological and pathophysiological mechanisms, and grouped together in the Obsessive-Compulsive and Related Disorders chapter of the DSM-5 (8); evidence supporting this nosological approach requires comparative studies between BDD and OCD. Dysfunctional connectivity in fronto-striatal networks is implicated in both OCD and BDD and overlaps with networks responsible for executive functions (EF) (9, 10). The function of these neural circuits can be assessed by specific psychometric tests (11, 12). Spatial working memory captures our ability to retain and organize visuospatial information, which involves frontoparietal circuitries (13, 14). Attentional set shifting, which is the ability to switch focus, is a form of cognitive flexibility that depends on the function of ventrolateral prefrontal cortex (15). Response inhibition, a measure of impulsivity, measured in the present study with the Stop Signal Task, is dependent on the inferior frontal cortex and its subcortical connections (11).

Studies using computer-standardized and validated neuropsychological tests such as the Cambridge Neuropsychological Test Automated Battery (CANTAB) have been used to assess important aspects of EF in patients with OCD and BDD. Three studies found that patients with OCD perform worse on the Spatial working memory task (SWM) compared with healthy controls and one additional study suggests that there could be a sex difference in task performance among patients with OCD (16-19). Similarly, Dunai et al. found worse performance on the SWM in patients with BDD compared with healthy controls (20). Worse performance on the Intra- extra dimensional set shifting (IED) in patients with OCD has been reported by Nedeljkovics et al., Watkins et al. Purcell et al., and Chamberlain et al. (16, 21-23), but not replicated by Purcell et al., Nielen et al., Veale et al., and Purcell et al. (17, 24, 25). Both Jefferies-Sewell et al., and Greenberg et al., showed that patients with BDD performed poorer on IED compared with healthy controls (26, 27). Stop signal task (SST) has been found to be deficient in patients with OCD in several studies (11, 18, 23, 28-31), but recent findings by Kalanthroff et al. could not demonstrate any difference, independent of medication status (32). Similarly, Jefferies-Sewell et al. showed worse test performance on the SST among patients with BDD in comparison to healthy controls (27). In summary, these studies suggest that similar EF could be deficient in patients with OCD and BDD, however, the results are inconclusive for OCD and scarce for BDD.

Direct comparisons between patients with OCD and BDD on EF are scarce. A study that compared 14 patients with BDD to 23 patients with OCD from previously published data detected similar, but not equivalent task performance on spatial span, spatial working memory, executive planning and pattern recognition (33). Another study showed poorer test performance on executive planning and the Stroop test between 14 patients with BDD and 10 patients with OCD compared to healthy controls (34). The question whether patients with OCD and BDD patients have similar or separate EF has important implications, since knowledge about neurocognitive performance in OCD and BDD could improve our neurobiological understanding and facilitate classification of these disorders, uncovering potentially important predictors of treatment outcome, and inform clinical care for these patients.

The aim of this study was therefore to use CANTAB to compare EF (spatial working memory, set shifting and response inhibition) between patients with OCD, BDD, and healthy controls. We hypothesized that OCD and BDD would exhibit similar executive dysfunction compared with healthy controls. Furthermore, we hypothesized that higher burden of symptoms is correlated with poorer EF among patients with OCD and BDD.

## METHODS

### Participants

Patients with OCD (n=29) were recruited from two psychiatric outpatient clinics in Stockholm specialized in obsessive-compulsive and related disorders. Patients with BDD (n=26) were participants in two studies on internet-delivered cognitive behavior therapy for BDD, conducted at Karolinska Institutet (35, 36). Inclusion criteria were a principal diagnosis of OCD or BDD according to the Diagnostic and Statistical Manual of Mental Disorders, fifth edition (DSM-5) (8). Exclusion criteria were i) psychotropic medication changes within at least two weeks prior to inclusion ii) completed cognitive behavioral therapy for OCD or BDD within the last 12 months, iii) comorbid OCD or BDD respectively, iv) current substance dependence or abuse, v) bipolar disorder, vi) psychotic disorders, vii) acute suicidal ideation, viii) severe personality disorder and ix) concurrent psychological treatment. Healthy controls (n=28) similar in age, sex and education, were recruited by online advertisement in the greater Stockholm catchment area. Exclusion criteria for healthy controls consisted of previous or present psychiatric disorder, psychotropic medication or chronic somatic disease. All participants were screened for eligibility and subsequently underwent a diagnostic interview using Mini International Neuropsychiatric Interview (M.I.N.I) and Structured clinical interview (SCID) with an experienced psychologist or psychiatrist before inclusion.

### Ethics

The regional ethical review board in Stockholm approved the study (registration ID 2015/1088-32 and 2013/1773-31/4). All trial subjects gave their written consent prior to participation in the study which adheres to the Declaration of Helsinki.

### Psychometric scales

Symptom severity of patients was assessed using the Yale-Brown Obsessive Compulsive Scale (YBOCS) for OCD or the modified version (BDD-YBOCS) for BDD (37, 38). Depressive symptoms were assessed with the self-rated version of the Montgomery-Åsberg Depression Rating Scale (MADRS-S) (39). The National Adult Reading Test, NART-SWE, was administered to all participants. NART-SWE is a validated test with sufficient reliability for assessment of premorbid IQ (40).

### Neurocognitive tests

Three tests from the Cambridge Neuropsychological Test Automated Battery (CANTAB) were administered in a fixed order and in a quiet room with a trained administrator giving standardized instructions. The tests were chosen to assess neurocognitive abilities of the participants, and key outcome measures for each test are presented below:

#### Spatial working memory

(SWM) measures the ability to retain and systematically organize visuospatial information (41). Participants are instructed to search for blue tokens hidden inside colored squares on the screen. Four levels of difficulty: three, four, six or eight boxes are presented at the same time. Two outcome measures are obtained. A between-search error means revisiting boxes in which a token has already been found. Strategy scores reflect how often search sequences are initiated from the same box within a trial.

#### Intra- extra dimensional set shifting

(IED) measures cognitive flexibility and specifically the ability to shift attention from irrelevant to relevant stimuli (42). Study subjects are asked to choose the right figure from two artificial dimensions with two figures each. Through trial and error, the rule of the game can be acquired. The rule changes after each block and at block eight the previously relevant dimension becomes irrelevant and vice versa. This is called the extra-dimensional shift, a measure of reversal learning. Outcome measures of interest include number of stages completed, total number of errors and number of errors in the extra dimensional shift.

#### The Stop Signal Task

(SST) measures response inhibition (43). Left and right arrows are presented randomly on a screen and participants are asked to press the corresponding button as fast as possible (go trial). However, participants are instructed to inhibit the response of pressing a button if an auditory signal is given, which occurs after an arrow is shown (no-go trial). The main outcome variable, Stop Signal Reaction Time (SSRT), is the mean time in milliseconds (ms) taken by the participant to suppress a prepotent response. The average reaction time for ‘go’ trials (i.e., trials without a stop signal) is a measure of a subject’s reaction time (psychomotor speed).

### Statistical analysis

Baseline demographic differences (age, sex, education, IQ, depressive symptoms and medication status) are presented as summary statistics, and between-group differences were tested using ANOVA for continuous variables and chi square for ordinal variables. Multiple linear regression models, controlled for age, sex, NART-SWE score and MADRS-S depressive symptoms, were used to compare test performance between the groups. Standardized effect sizes (Cohen’s *d*) were obtained by dividing the estimated marginal means by the residual degrees of freedom from the regression model described above, using the standard deviation of the model residuals as sigma (population standard deviation) as implemented in the *emmeans* R package (44). For the two clinical groups, symptom severity scores (Y-BOCS and BDD-YBOCS, respectively) were standardized and compared to test performance using main diagnosis as covariate. Pearson’s *r* was used as a measure of effect size. The linear models were deemed appropriate for the data by inspecting residuals, homoscedasticity and outliers using diagnostic plots. Missing values (<10% for all variables) were assumed to be missing at random (MAR) and were imputed using bagged trees, a non-linear ensemble method that can impute both continuous and categorical data. In bagged trees imputation, each missing value is predicted in bootstrap resampling of data and aggregated to form a single prediction using averages for continuous values and majority vote for categorical outcomes. Regular bagged trees allow for non-linear ensemble imputation whereby all predictors are available, and the best one is selected, at each decision node split for each tree in the ensemble (45, 46). All statistical analyses were performed using R version 4.0.2 and STATA version 15 (StataCorp: College Station)(47). Code for the statistical analyses is available on the Open Science Framework (https://doi.org/10.17605/OSF.IO/63YNF).

## RESULTS

### Demographic and clinical characteristics

In the present sample, groups did not differ in age, sex, or highest completed education (Table 1). The proportion of patients on SSRI was higher in the OCD group compared with the BDD group, and patients with OCD had lower NART-SWE premorbid IQ. Moreover, OCD and BDD groups scored higher on depressive symptoms compared to healthy controls. On average, symptom severity was in the moderate-to-severe range for OCD and BDD groups. See **Table 1** for demographic and clinical characteristics of the participants.

**Table 1.** Sociodemographic and clinical characteristics of participants with Obsessive-compulsive disorder, Body Dysmorphic disorder and healthy controls

| Demographic variables, n (%) | Control (n = 28) | OCD (n = 29) | BDD (n = 26) | Missing values, n (%) |
| --- | --- | --- | --- | --- |
| Age, Mean (SD) | 30.8 (11.9) | 31.2 (9.2) | 30.7 (9.5) | 1 (1%) |
| Sex; Female count (%) | 19 (68%) | 15 (52%) | 18 (70%) | 1 (1%) |
| Highest completed education; % |  |  |  | 0 |
| Primary school | 1 (3.6%) | 2 (6.9%) | 0 |  |
| Vocational school | 1 (3.6%) | 2 (6.9%) | 0 |  |
| High school | 16 (57%) | 14 (48.2%) | 18 (69.2%) |  |
| University degree | 10 (35.7%) | 10 (34.5%) | 7 (26.9%) |  |
| Ph.D | 0 | 1 (3.4%) | 1 (3.85%) |  |
| NART-SWE | 37.9 (5.0) | 33.4 (5.8) * | 37.7 (5.8) | 3 (4%) |
| MADRS score (0-56), mean (SD) | 2.8 (2.6) | 21.4 (8.8) * | 17.1 (8.0) * | 1 (1%) |
| Clinical variables (OCD & BDD group only) |  |  |  |  |
| YBOCS score, mean, (SD) |  | 24.9 (5.2) | N/A | 0 |
| BDD-YBOCS score, mean (SD) |  | N/A | 29,4 (5,4) | 0 |
| SSRI medication |  | 23 (79%) * | 5 (19%) * | 0 |
| Comorbid conditions |  |  |  | 5 (9%) |
| Depression |  | 10 (37%) | 13 (57%) |  |
| Social phobia |  | 2 (7%) | 5 (22%) |  |
| Panic disorder |  | 2 (7%) | 1 (4%) |  |
| Generalized anxiety disorder |  | 2 (7%) | 1 (4%) |  |
| Bulimia nervosa |  | 0 | 1 (4%) |  |
| Tourette's syndrome |  | 3 (11%) | 0 |  |
| Trichotillomania |  | 1 (4%) | 0 |  |

OCD, Obsessive-compulsive disorder; BDD, Body dysmorphic disorder; Nart-Swe, National adult reading test-
estimated verbal IQ in Swedish; MADRS, Montgomery-Åsberg Depression Rating Scale; Y-BOCS, Yale Brown Obsessive Compulsive Scale; BDD-YBOCS, Yale Brown Obsessive Compulsive Scale for Body Dysmorphic Disorder, SSRI, Selective serotonin receptor inhibitors. \* P-value <0.05.

### Neurocognitive task performance

Table 2 shows estimated means and standardized effect sizes across groups on the specific CANTAB tests.

**Table 2.** Estimated mean group differences on principal outcomes for Spatial working memory, Intra- extra dimensions set shifting and Stop signal task

| Group, Estimated mean (SE) |  |  |  | Group comparisons, Cohen's *d* [95% confidence interval] |  |  |
| --- | --- | --- | --- | --- | --- | --- |
|  | Control | OCD | BDD | Control - OCD | Control - BDD | BDD - OCD |
| **Spatial Working Memory** |  |  |  |  |  |  |
| Total between-search errors | 15.22 (4.28) | 23.32 (3.73) | 21.4 (3.40) | -0.51 [-1.35 to 0.34] | -0.39 [-1.10 to 0.32] | -0.12 [-0.73 to 0.49] |
| Strategy score | 30.06 (1.54) | 31.47 (1.34) | 31.90 (1.22) | -0.24 [-1.08 to 0.59] | -0.32 [-1.03 to 0.39] | 0.07 [-0.53 to 0.68] |
| **Intra- extra Dimensional set shifting** |  |  |  |  |  |  |
| Total errors | 16.89 (3.25) | 21.13 (2.84) | 17.47 (2.58) | -0.35 [-1.19 to 0.49] | -0.05 [-0.75 to 0.66] | -0.30 [-0.91 to 0.31] |
| Extra dimensional errors | 7.82 (2.55) | 10.11 (2.23) | 9.23 (2.03) | -0.24 [-1.08 to 0.60] | -0.15 [-0.86 to 0.56] | -0.09 [-0.70 to 0.51] |
| **Stop Signal Task** |  |  |  |  |  |  |
| Stop signal reaction time (ms) | 216.59 (30.4) | 224.53 (26.5) | 241.58 (24.16) | -0.07 [-0.91 to 0.77] | -0.22 [-0.93 to 0.49] | 0.15 [-0.46 to 0.76] |
| Mean reaction time (ms) | 428.99 (27.88) | 410.26 (24.3) | 445.9 (22.15) | 0.18 [-0.66 to 1.02] | -0.16 [-0.87 to 0.55] | 0.34 [-0.27 to 0.95] |

Abbreviations: BDD, Body Dysmorphic disorder; OCD, Obsessive-compulsive disorder; SE, standard error.
Estimated marginal means based on the linear models using age, sex, language score and depressive symptoms as covariates.

### Spatial working memory

There were no statistically significant differences in SWM strategy score when comparing the OCD group (Estimate = 1.41 [95% CI -3.42 to 6.24], SE = 2.43, *p* = 0.56) and BDD group (Estimate = 1.84 [95% CI -2.24 to 5.92], SE = 2.05, *p* = 0.37) to healthy controls. Further, there were no statistically significant main effects of age, sex, NART-SWE score, or MADRS-S score.

Similarly, for the SWM between errors estimates for each group, there were no statistically significant differences between the OCD group (Estimate = 8.11 [95% CI -5.30 to 21.52], SE = 6.74, *p* = 0.23) and BDD group (Estimate = 6.18 [95% CI -5.14 to 17.50], SE = 5.68, *p* = 0.28). There was, however, a small but statistically significant main effect of age (Estimate = 0.41 [95% CI 0.04 to 0.78], SE = 0.19, *p* = 0.03) showing an association between higher age and a higher SWM between errors score.

### Intra- extra dimensional set shifting

All participants in the groups passed stages 1-7. Twenty-four percent (n=7) of the OCD group failed to pass the extra-dimensional shift stage (EDS stage, stage 8) compared to 19% (n=5) in the BDD group and 10% (n=3) of the control subjects. Neither the OCD group (Estimate = 4.23 [95% CI -5.97 to 14.43], SE = 5.12, *p* = 0.41) nor the BDD group (Estimate = 0.57 [95% CI -8.04 to 9.18], SE = 4.32, *p* = 0.89) differed from the control group in total IED errors, however there was a statistically significant effect of sex where women had a higher mean than men (Estimate = 6.11 [95% CI 0.32 to 11.90], SE = 2.91, *p* = 0.04).

Extra-dimensional shift errors were similar with no differences between the OCD (Estimate = 2.29 [95% CI -5.72 to 10.30], SE = 4.02, *p* = 0.57) and BDD (Estimate = 1.41 [95% CI -5.35 to 8.17], SE = 3.40, *p* = 0.68) groups compared to healthy controls, but a statistically significant effect of sex where females scored higher (Estimate = 8.64 [95% CI 4.10 to 13.19], SE = 2.28, *p* < 0.001).

### Stop signal task

The difference in Stop signal reaction time did not differ between the OCD (Estimate = 7.94 [95% CI -87.41 to 103.29], SE = 47.88, *p* = 0.87) and BDD (Estimate = 24.99 [95% CI -55.50 to 105.48], SE = 40.42, *p* = 0.54) groups compared to healthy controls, and there were no statistically significant associations with other covariates.

Similarly, the mean reaction time did not differ between the OCD group (Estimate = -18.72 [95% CI -106.16 to 68.71], SE = 43.91, *p* = 0.67) and the BDD group (Estimate = 16.91 [95% CI -56.89 to 90.72], SE = 37.07, *p* = 0.65) compared to healthy controls, however there was a statistically significant main effect of age (Estimate = 3.06 [95% CI 0.64 to 5.47], SE = 1.21, *p* = 0.01) where a higher age was associated with a longer average reaction time.

### Symptom severity and neurocognitive task performance

In a subgroup analysis of the patients with OCD and BDD we investigated a standardized symptom severity score based on the Y-BOCS and BDD-YBOCS total scores, respectively, as well as group. Figure 1 displays the correlations between symptom severity and neurocognitive task performance.

**Figure 1.**
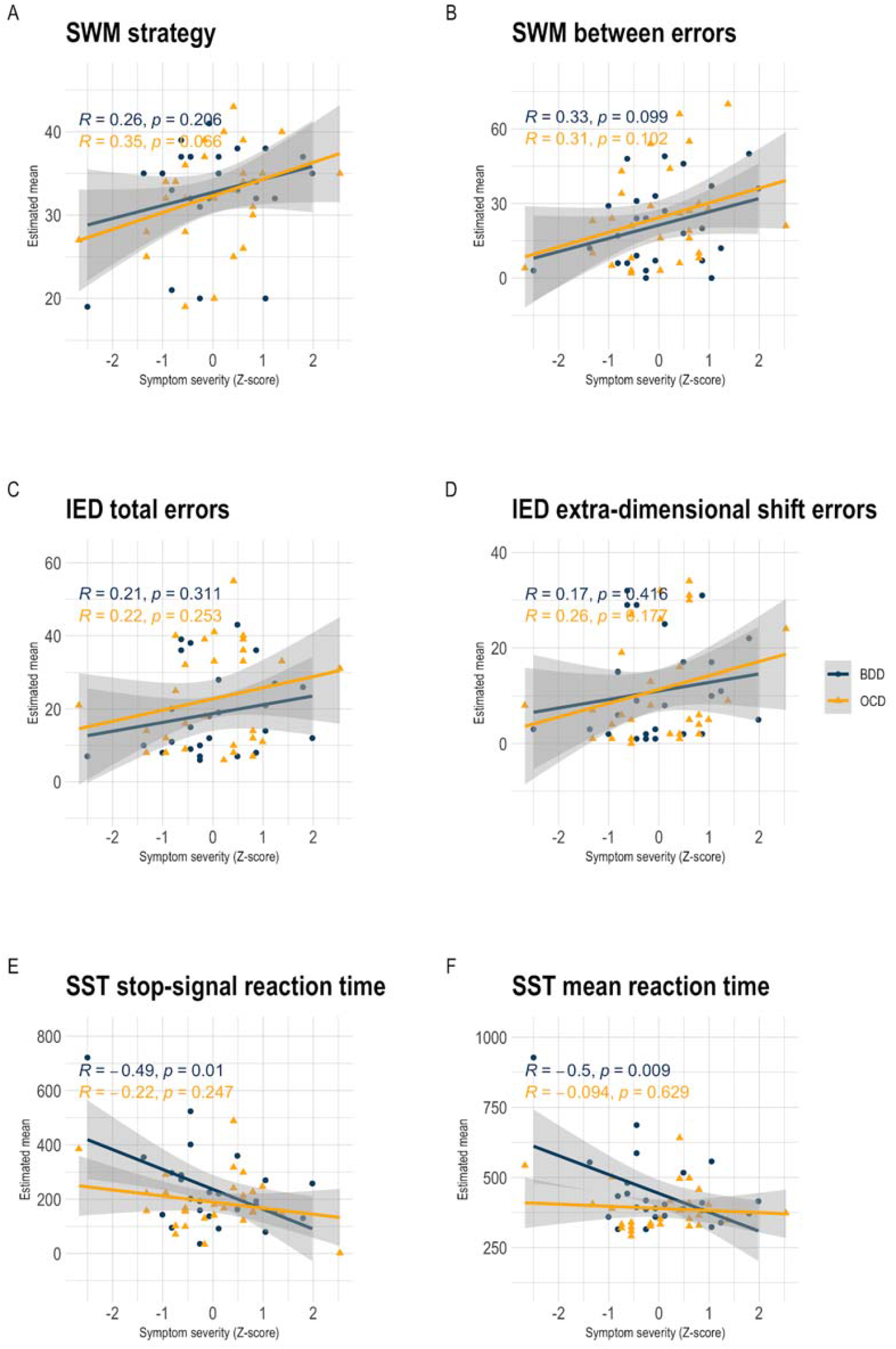
Linear associations between symptom severity and executive function in OCD and BDD clinical groups.

### Spatial working memory

Symptom severity was statistically significantly associated with SWM strategy (Estimate = 1.80 [95% CI 0.22 to 3.78], SE = 0.79, *p* = 0.03), and there was a positive but not statistically significant correlation between symptom severity and SWM strategy score (OCD: *r* = 0.35, *p* = 0.066; BDD: *r* = 0.26, *p* = 0.206).

The model for SWM between errors showed an association between symptom severity and test performance (Estimate = 5.64 [95% CI 0.95 to 10.33], SE = 2.41, *p* = 0.02), which was also seen in the correlation between symptom severity and test performance in the OCD (*r* = 0.31, *p* = 0.102) and BDD (*r* = 0.33, *p* = 0.099) groups.

### Intra- extra dimensional set shifting

There was no statistically significant association between symptom severity and intra- extra dimensional total errors (Estimate = 2.76 [95% CI -0.76 to 6.28], SE = 1.75, *p* = 0.12), with weak correlations between symptom severity and performance in both the OCD (*r* = 0.22, *p* = 0.253) and BDD (*r* = 0.21, *p* = 0.311) groups.

Similar results were found in the analysis of extra dimensional errors, with no statistically significant association between symptom severity (Estimate = 2.37 [95% CI -0.62 to 5.36], SE = 1.59, *p* = 0.12), and weak correlations in both groups (OCD: *r* = 0.26, *p* = 0.177; BDD: *r* = 0.17, *p* = 0.416).

### Stop signal task

The model for Stop signal reaction time indicated a statistically significant association between symptom severity and task performance (Estimate = -46.31 [95% CI -78.69 to -13.95], SE = 16.13, *p* = 0.005), and correlations differed between the groups with a statistically significant correlation in patients with BDD (*r* = -0.49, *p* = 0.01) but not in patients with OCD (*r* = -0.22, *p* = 0.247). When analyzing mean reaction time, there was an effect of symptom severity (Estimate = -35.76 [95% CI -64.49 to -7.04], SE =14.31, *p* = 0.02) and a moderate negative correlation between symptom severity and mean reaction time in patients with BDD (*r* = -0.5, *p* = 0.009) but not OCD (*r* = -0.094, *p* = 0.629).

### Sensitivity analyses

Results were attenuated but similar when adding age, sex, and SSRI medication status into the models. Both SWM strategy score (Estimate = 1.81 [95% CI 0.01 to 3.61], SE = 0.90, *p* = 0.05) and between errors (Estimate = 8.71 [95% CI 3.56 to 13.85], SE = 2.56, *p* = 0.001) showed an association with clinical symptoms. As in the main analysis, symptom severity showed a weak association to both IED total errors (Estimate = 2.37 [95% CI -1.73 to 6.47], SE = 2.04, *p* = 0.25) and IED extra dimensional errors (Estimate = 0.84 [95% CI -2.35 to 4.04], SE = 1.59, *p* = 0.60). Finally, the estimates from the stop signal reaction time (Estimate = -46.45 [95% CI -81.59 to -11.31], SE = 17.49, *p* = 0.01) and mean reaction time (Estimate = -30.37 [95% CI -61.58 to 0.84], SE = 15.53, *p* = 0.06) were similar but slightly weakened. Overall, associations between clinical symptoms and CANTAB test scores were in the same direction and of similar magnitude in the sensitivity analyses compared to main analyses.

## DISCUSSION

In this study, we compared executive functions (EF) between patients with OCD, BDD and healthy controls using computerized and standardized tests. There were no statistically significant differences in group means for either of the three tests: Spatial working memory (SWM), Intra- extra dimensional set-shifting (IED), and Stop signal task (SST). Age was found to be a covariate associated with performance on SWM and SST, whereas sex was associated with performance on IED. There were statistically significant associations between symptom severity and test performance in SWM between errors and the SST, but not for the IED task. The correlation coefficients indicated that higher symptom severity was associated with worse performance for both the BDD and OCD groups in SWM, but reverse correlation for the BDD group in the SST.

Our findings differ from previous studies in that the clinical groups did not perform significantly worse than healthy controls. One explanation could be that the clinical cases in previous studies were more severe than the present cases, yet this seems unlikely given that the present cases were regular psychiatry patients diagnosed, assessed and treated by gold standard procedure. Instead, comparing crude numbers on performance on IED and SST shows that patients with OCD and BDD in our study are comparable to those in previous studies, however, healthy controls perform much better in previous studies (26, 27). Recruiting healthy controls from universities constitutes a risk for selection bias, since university students may have prominent EF and are therefore not representative for the general population. In addition, OCD is a prevalent comorbidity in patients with BDD from previous studies, up to 75 % of all participants in one study (27). The evidence for deficient EF in all three tests is more robust in patients with OCD compared with studies on BDD (48, 49). Therefore, high comorbidity of OCD in a BDD population could be a major confounding factor when interpreting the results.

It has been suggested that certain executive dysfunctions, such as cognitive inflexibility and deficient response inhibition, constitute an increased risk for developing compulsive behaviors, due to inadequate habit formations, inability to stop a response or sequence, and failure to maintain goal-directed behavior (50-52). Nevertheless, using neuropsychological tests to probe for specific EF is treacherous since EFs require multiple cognitive processes (53). Especially considering that comparability between studies is limited by the use of different neuropsychological tests, both manually administered and computer-standardized tests (54).

Even if patients with OCD and BDD had impaired EF in cognitive flexibility or response inhibition, other cognitive processes could compensate for substantial abnormalities, which has been suggested for IED and SST in patients with OCD (55-57). Moreover, major psychiatric disorders, such as attention deficit hyperactivity disorder, major depressive disorder, bipolar disorder, and schizophrenia, are associated with deficits in EF with similar or larger effect sizes (58-60). This is in line with meta-analyses of patients with OCD showing broad impairment in EF, but small to moderate effect sizes compared to healthy controls (48, 49, 61). This raises the question whether specific executive dysfunctions are characteristic for patients with OCD and BDD or whether they exhibit nonspecific cognitive impairments.

Symptom severity was associated with worse performance on some, but not all, tasks in the CANTAB for both diagnostic groups. This is in line with previous literature, where results are inconsistent. Furthermore, methodological shortcomings from previous studies limits the generalizability of these findings (62). The lack of a clear association between symptom severity and neuropsychological test-results dampens expectations that results from neurocognitive tests can serve as markers of severity or predictors of treatment outcomes. However, impaired EF has been suggested to impact the outcome of cognitive behavioral therapy (63-65) and well powered longitudinal studies are warranted to further study these associations.

## Limitations

Firstly, EF involve several higher-order cognitive processes and the tasks that participants have accomplished in this study assess important aspects of it. However, for comprehensive characterization of EF in patients with obsessive-compulsive and related disorders, other neuropsychological tests could be considered (66). Secondly, patients with OCD had significantly lower NART-SWE scores and used medication to a greater extent than patients with BDD or healthy controls, although previous studies do not suggest a correlation between IQ and EF (27, 28). Moreover, studies so far have not yielded convincing results that indicate significant interaction between serotonin reuptake inhibitor (SSRI) and cognitive functions (24, 32). Lastly, a small sample size can limit the power to detect significant differences (67). However, despite a comparable or larger sample size compared with previous studies, we did not see a generally deficient EF in patients with OCD or BDD.

## Conclusion

Patients with OCD, BDD, and healthy controls performed comparably on neuropsychological tests assessing EF, tentatively suggesting that EF in spatial working memory, cognitive flexibility and response inhibition are similar on a group level. There were associations between symptom severity and test performance on some, but not all, neurocognitive tests for the participants with OCD and BDD, generally indicating worse performance with higher symptom severity. A larger sample size might be needed to determine the presence or absence of group differences in EF with more certainty, especially in patients with BDD. Moreover, longitudinal studies may reveal the clinical relevance of neuropsychological test performance for prognosis and treatment outcome.

## Data Availability

Code for the statistical analyses is available on the Open Science Framework (https://doi.org/10.17605/OSF.IO/63YNF).

